# The Way Home: A Scoping Review of Public Health Strategies to Increase the Utilization of Home Dialysis in Chronic Kidney Disease Patients

**DOI:** 10.1101/2024.12.29.24319745

**Authors:** Natcha Yongphiphatwong, Yot Teerawattananon, Pitsinee Supapol, Denla Pandejpong, Tanainan Chuanchaiyakul, Jiratorn Sutawong, Naline Gandhi, Nutthawongse Kiatkrissada, Saudamini Vishwanath Dabak, Thunyarat Anothaisintawee

**Author notes:** Corresponding author, Address: Faculty of Medicine, Ramathibodi Hospital, Mahidol University, Bangkok, Thailand. These authors made equal contributions (Pitsinee Supapol, Denla Pandejpong, Tanainan Chuanchaiyakul, Jiratorn Sutawong, Naline Gandhi).

## Abstract

**Introduction:** Home dialysis (HoD) remains underutilized, despite evidence showing it provides comparable mortality rates to in-center hemodialysis (ICHD) while offering advantages such as improved quality of life and lower overall costs. This scoping review comprehensively evaluates the impact of public health interventions on increasing the use of HoD, including both Peritoneal Dialysis (PD) and Home Hemodialysis (HHD).

**Methods:** Relevant studies were searched in the Web of Science, Medline, Embase, Scopus, EBSCOhost, and EconLit databases from their inception through May 2024. Studies were eligible for review if they assessed the effectiveness of public health interventions in terms of utilization and retention rates for general HoD, PD, and HHD.

**Results:** Forty-three studies were included, with interventions categorized into three main types: educational programs, service provision improvements, and modifications to payment structures. Our findings indicate that educational interventions—aimed at enhancing knowledge about dialysis options and promoting shared decision-making among patients, families, and healthcare providers—and service provision improvements, such as assisted PD and nephrologist-performed catheter insertions, could significantly increase the initiation, utilization, and retention rates of HoD. However, the impact of payment interventions on HoD outcomes differed across different contexts.

**Conclusion:** Education and service provision enhancements may represent the most effective public health interventions for increasing initiation, utilization, and retention rates of HoD in dialysis requiring patients. However, these findings are predominantly based on evidence from observational studies; further experimental studies with rigorous methodology are warranted to validate the effectiveness of these interventions in promoting HoD utilization.

**PLAIN TEXT SUMMARY:** Kidney dialysis is a life-sustaining therapy that can be offered both at home and in medical centres, however, home dialysis is underutilised globally. This scoping review gathers evidence from around the world to identify and assess the effectiveness of public health interventions to improve home dialysis utilization. The interventions we found were mainly related to improving patient knowledge, redesigning service provision, or adjusting payment/reimbursement conditions. Our results suggest that educating patients about their dialysis options to support shared decision-making before they require dialysis and offering assisted peritoneal dialysis at home can help increase the number of patients starting and staying on home dialysis. However, adjusting payment and reimbursement policies showed mixed results.

## INTRODUCTION

Chronic Kidney Disease (CKD) is a significant public health burden with a global prevalence of 13.4% (11.7-15.1%)^1^. CKD is associated with an increased risk of all-cause and cardiovascular mortality alongside substantial impacts on patients’ daily lives. As kidney function declines, health-related quality of life (HRQoL) deteriorates, affecting the physical, emotional, and social well-being of patients^2,3^. According to the Kidney Outcomes Quality Initiative (KDOQI) guideline^4^, CKD can be classified into five stages using the estimated glomerular function (eGFR) parameter and/or evidence of structural renal changes e.g. proteinuria. CKD stage 5 is defined as eGFR of less than 15 mL/min/1.73 m^2^ and is also referred to as end-stage kidney disease (ESKD). At this stage patients typically require Kidney Replacement Therapy (KRT) including kidney transplantation (KT) or dialysis –to replace lost kidney function. While KT yields the highest HRQoL among available treatment options, access to KT is limited due to a shortage of both living and cadaveric donors^5–8^. Consequently, most ESKD patients depend on dialysis for ongoing care.

Dialysis options include in-centre hemodialysis (ICHD), peritoneal dialysis (PD), and home hemodialysis (HHD), with the latter two offering the flexibility of home-based care, meaning that they can be carried out by patients or their caregivers in the comfort of their homes. Findings from systematic reviews and meta-analyses suggest that PD had comparable mortality risk to ICHD^9^. Additionally, PD patients experience fewer cardiovascular events^10^ and report a better health-related quality of life (HRQoL) compared to those on ICHD^11,12^. In terms of value for money, evidence from high-income countries (HICs) indicates that PD is more cost-effective than ICHD^7,13–15^. Moreover, in low- and middle-income countries (LMICs), a cost-effectiveness analysis conducted in Thailand also found that when compared to palliative care, the average incremental cost-effectiveness ratio (ICER) for initial treatment with PD was lower than that for ICHD^16^.

Although PD is associated with lower costs and improved patient HRQoL compared to ICHD, it remains significantly underutilized, particularly in LMICs^17^. A global survey highlighted the disparity, revealing that the utilization of PD in low-income countries (LICs) is 60 times lower than in HICs, with PD use at just 0.9 per million population (pmp) (95% CI: 0.7–1.5) in LICs, compared to 53.0 pmp (95% CI: 40.6–89.8) in HICs^17^. Several barriers limit the utilization of PD in both HICs and LMICs. These include insufficient education on the available KRT options, leading to a lack of shared decision-making between patients and healthcare providers^18^. Additionally, inadequate support for PD services—such as limited PD expertise and insufficient clinical training for physicians and nurses^19^—low provider reimbursement^20^, and unsuitable home environments for PD^21^ further hinder its use. These barriers pose challenges to both international and local recommendations aimed at enhancing home-based treatments for dialysis-requiring patients.

In Thailand, PD utilization declined dramatically, following the 2022 shift from the “PD-First” policy to one where patients may select their preferred dialysis modality. While the new policy offers greater autonomy to patients, its aftermath included lowered ICHD quality due to service capacity overload, a sharp increase in the dialysis budget, and a severely threatened PD ecosystem due to reduced patient volumes^22,23^. In an effort to mitigate these effects, a government-commissioned working group (WG) in Thailand has recommended increasing PD utilization from 15% to 50%. Therefore, to inform the WG, we conducted a scoping review of the effectiveness of public health interventions in increasing the utilization of HoD, including both PD and HHD. This scoping review aims to assess the effects of the interventions on the uptake and retention of HoD utilization across the world. HHD was included in this review as the lessons learned from HHD provision in HICs may also apply to PD provision in lower- and middle-income contexts. Beyond informing the WG, the findings of this review can also provide valuable insights for the global kidney community.

## RESULTS

A comprehensive search yielded 25,067 studies, as shown in Figure 1. After removing 7,774 duplicates, the title and abstract of the remaining 17,283 studies were screened, resulting in 726 studies whose full texts were assessed for eligibility. Of the 726 full texts assessed, 42 studies met the inclusion criteria, and a thorough review of the reference lists of the selected studies further identified one additional study. Thus, 43 studies were included in this scoping review^24–66^.

**Figure 1.**
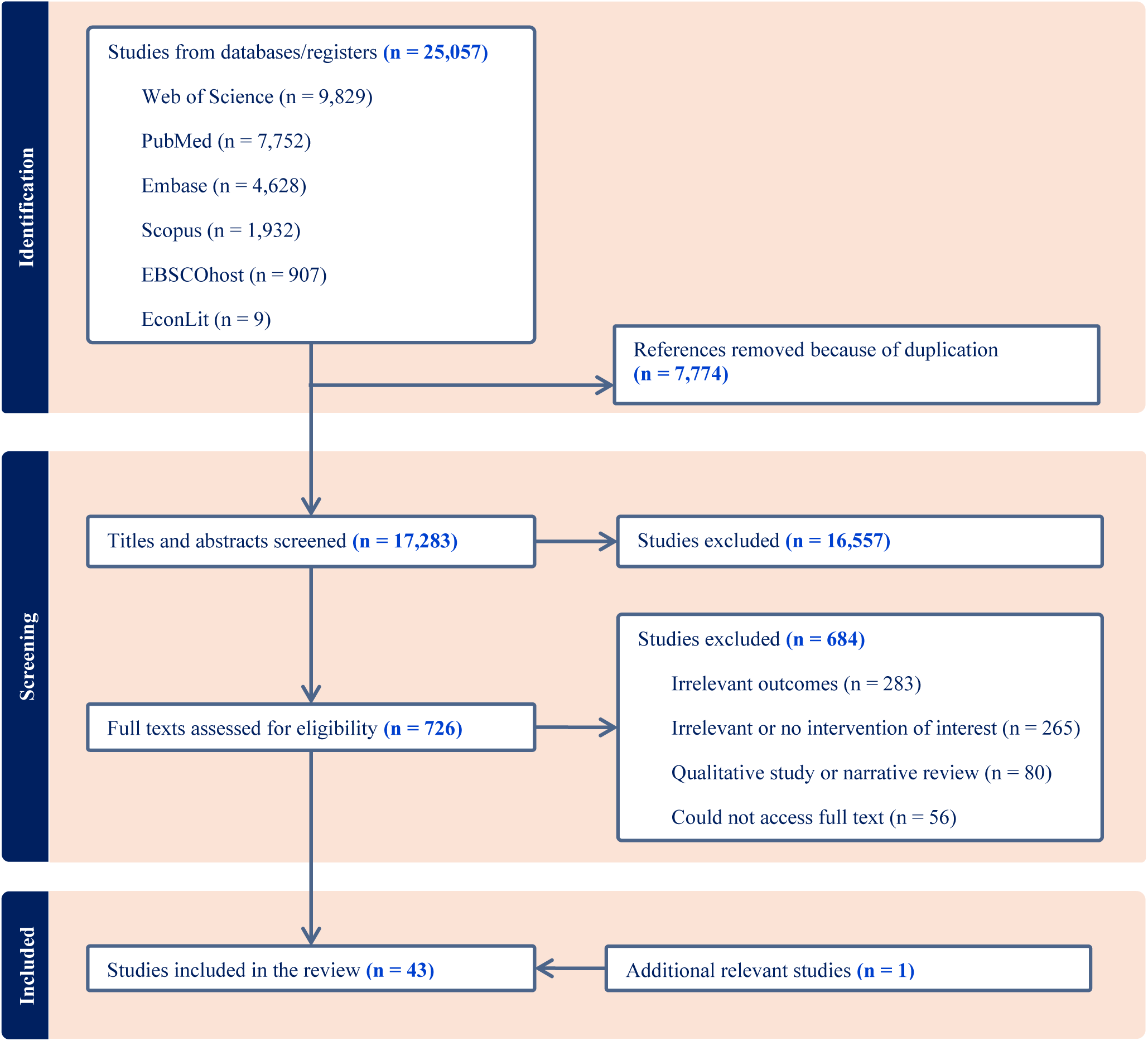
PRISMA Diagram.

Characteristics of the included studies are presented in Table 1. All the included studies were published within the last two decades with 49% of included studies published from 2020 onwards^34–40,48–55,61–66^, and 42% published in the 2010s^24–33,44–47,57–60^. The majority of studies were conducted in the Region of the Americas (AMR) and Western Pacific Region (WPR) according to the World Health Organization regions, with 15 of 43 (35%) studies conducted in the United States of America (USA)^25,30,31,33,36,37,39,41,53,58–61,64,66^. Additionally, 39 of the 43 included studies (91%) were conducted in HICs^24–33,35–43,45,47–56,58–66^, as defined by the World Bank’s income group, while the remaining four studies were conducted in upper-middle-income countries (i.e., China^44,46^ and Thailand^34,57^).

**Table 1.** Characteristics of included studies.

| First Author<br>(Year) | Study Design | Country/<br>Region | Study<br>Setting | Intervention | Comparator | Intervention<br>Lead | Target<br>Population |
| --- | --- | --- | --- | --- | --- | --- | --- |
| *Education* |  |  |  |  |  |  |  |
| Castledine (2013) | Retrospective Cohort | UK | National Level | Pre-dialysis education program using home visit | No intervention | Nurses and Existing Patients | Patients |
|  |  |  |  | Pre-dialysis education program using group session |  |  |  |
|  |  |  |  | Pre-dialysis education program using Existing Patients |  |  |  |
|  |  |  |  | Pre-dialysis education program using review of modality |  |  |  |
|  |  |  |  | Pre-dialysis education program using video/DVD materials |  |  |  |
| Tamura (2013) | Case-Control | USA | National Level | Kidney Early Evaluation Program (KEEP) for screening and education | No intervention | Healthcare Providers | Patients |
| Fortnum (2014) | Cross-sectional | Australia | National Level | Renal units offering more group sessions per year | No intervention | Healthcare Providers | Patients |
| Chan (2015) | Retrospective Cohort | Canada | Single Center | Simulation-based teaching | Conventional teaching | Nurses | Patients |
| Prieto-Velasco (2015) | Prospective Cohort | Spain | Multiple Centers | Education Process (EP) with Patient Decision Aid (PDA) tools | No intervention | Multidisciplinary Care Team | Patients |
| de Maar (2016) | Retrospective Cohort | Netherlands | Single Center | Pre-dialysis programme (GUIDE) | Historical control | Multidisciplinary Care Team | Patients |
| Shukla (2017) | Retrospective Cohort | USA | Single Center | Comprehensive pre-dialysis education program | United States Renal Data System | Multidisciplinary Care Team | Patients |
| Dubin (2019) | Prospective Cohort | USA | Multiple Centers | Digital Modality Decision Program | Historical control | Multidisciplinary Care Team | Patients |
| Lee (2019) | Prospective Cohort | Taiwan | Not Reported | Shared decision-making (SDM) | Historical control | Multidisciplinary Care Team | Patients |
| Shukla (2019) | Prospective Cohort | USA | Single Center | Comprehensive pre-dialysis education program | CKD care in USRDS data | Nurses and Nephrologists | Patients and Caregivers |
|  |  |  |  |  | Historical control |  |  |
| Parapiboon (2020) | RCT | Thailand | Single Center | Customized multimedia | Conventional multimedia | Dialysis Facilities | Patients |
| Imamura (2021) | Retrospective | Japan | Single | Multidisciplinary care (MDC) | No intervention | Multidisciplinary | Patients |

| First Author (Year) | Study Design | Country/Region | Study Setting | Intervention | Comparator | Intervention Lead | Target Population |
| --- | --- | --- | --- | --- | --- | --- | --- |
|  | Cohort |  | Center |  |  | Care Team |  |
| Shukla (2021) | Retrospective Cohort | USA | National Level | Kidney Disease Education (KDE) | No intervention | Government Healthcare Payers and Dialysis Facilities | Dialysis Facilities and Patients |
| McKeon (2022) | Retrospective Cohort | USA | Multiple Centers | A structured CKD Education Program | No intervention | Multidisciplinary Care Team | Patients |
| Shah (2022) | Prospective Cohort | UK | Single Center | Quality Improvement (QI) by training nurses and patients | Pre-QI Period | Healthcare Providers and Existing Patients | Nurses and Patients |
| Blankenship (2023) | Retrospective Cohort | USA | Multiple Centers | Transitional care units (TCUs) or dedicated care programs or dialysis orientation units | Historical control | Healthcare Providers | Patients |
| Sakurada (2023) | Retrospective Cohort | Japan | Single Center | Shared decision-making (SDM) | No intervention | Nephrologists and Nurses | Patients and Caregivers |
| *Service Provision* |  |  |  |  |  |  |  |
| Asif (2005) | Retrospective Cohort | USA | Multiple Centers | PD catheter insertion by nephrologists | PD catheter insertion by surgeon | Nephrologists | Patients |
| Oliver (2007) | Retrospective Cohort | Canada | Multiple Centers | Home Plus Program | No intervention | Nurses | Patients |
| Jiang (2011) | Cross-sectional | China | Multiple Centers | PD satellite center program | Historical control | Physicians and Nurses | Physicians and Nurses |
| Chen (2012) | Prospective Cohort | Taiwan | Multiple Centers | Multidisciplinary care (MDC) | Usual care group | Multidisciplinary Care Team | Patients |
| Castledine (2013) | Retrospective Cohort | UK | National Level | Provision home visits to PD patients | No intervention | Nurses and Existing Patients | Patients |
|  |  |  |  | PD catheter insertion by members of renal team | PD catheter insertion by surgeon |  |  |
|  |  |  |  | Provision same day hospital visits for PD patients | No intervention |  |  |
| Yu (2014) | Retrospective Cohort | China | Single Center | Continuous quality improvement | Historical control | Multidisciplinary Care Team | Patients |
| Blaauw (2019) | Prospective Cohort | UK | Not Reported | Remote patient management (RPM) systems | Historical control | Nurses | Nurses and Patients |
| Boyer (2020) | Retrospective | France | National | Nurse-assisted PD | Historical control | Nurses | Dialysis |
|  | Cohort |  | Level |  |  |  | Facilities and Patients |
| Liu (2021) | Retrospective Cohort | Singapore | Single Center | PD catheter insertion by nephrologists | Historical control | Nephrologists | Patients |
| van Eck van der Sluijs (2021) | Cross-sectional | Europe | Multiple Centers | Assisted PD program | No intervention | Nurses and Caregiver | Patients |
| Yao (2021) | Retrospective Cohort | Taiwan | National Level | PD center volume (26-42 incident patients per year) | PD center volume (1-12 incident patients per year) | Dialysis Facilities | Patients |
| Quinn (2024) | Prospective Cohort | Canada | Sub-National Level | At Home, on the Right Therapy (START) project | Historical control | Dialysis Facilities | Patients |
| **Combined Education and Service Provision** |  |  |  |  |  |  |  |
| Kaiser (2020) | Prospective Cohort | USA | Single Center | Virtual Multidisciplinary Care Program | No intervention | Multidisciplinary Care Team | Patients |
|  |  |  |  |  | Self-control (pre-education) |  |  |
| Tombocon (2021) | Prospective Cohort | Australia | Multiple Centers | Quality Improvement through establishing treatment pathways that coordinates local home treatment, raise awareness of HoD, and develop flexible individualized treatment (Home before Hospital) | Historical control | Multidisciplinary Care Team | Patients |
| Manns (2022) | RCT | Canada | Sub-National Level | Multifaceted Interventions, including phone surveys from a knowledge translation broker, 1-year center-specific audit/feedback on home dialysis use, delivery of an educational package, and an academic detailing visit | No intervention | Nephrologists | Dialysis Facilities |
| **Payment** |  |  |  |  |  |  |  |
| Mendelssohn (2004) | Case-Control | Canada | Sub-National Level | Equal physician reimbursement for all dialysis modalities | Historical control | Government Healthcare Payers | Physicians |
| Praditpornsilpa (2011) | Retrospective Cohort | Thailand | National Level | PD-first policy (2009) | Historical control (2007) | Government Healthcare Payers | Dialysis Facilities |
| Hirth (2013) | Retrospective | USA | National | Medicare Prospective Payment | Historical control | Government | Dialysis |
|  | Cohort |  | Level | System (PPS) | (2007) | Healthcare Payers | Facilities |
| Lin (2017) | Retrospective Cohort | USA | National Level | Medicare Prospective Payment System (PPS) | Historical control (2007) | Government Healthcare Payers | Dialysis Facilities |
|  |  |  |  | Add-on paying for home dialysis training (Medicare Parts A/B subgroup) | No intervention |  |  |
| Sloan (2019) | Retrospective Cohort | USA | National Level | Medicare Prospective Payment System (PPS) | Historical control | Government Healthcare Payers | Dialysis Facilities |
| Lin (2020) | Retrospective Cohort | USA | National Level | PD catheter paid for by Medicare | No intervention | Government Healthcare Payers | Patients |
| Sriravindrarajah (2020) | Retrospective Cohort | Australia | Sub-National Level | Private health insurance (PHI) | No intervention | Patients | Patients |
| Trachtenberg (2020) | Retrospective Cohort | Canada | Sub-National Level | Equal nephrologist fee-for-service (FFS) for HD and PD | Salaried nephrologist | Government Healthcare Payers | Physicians |
| Ji (2022) | RCT | USA | National Level | End-Stage Renal Disease Treatment Choices (ETC) Payment Model | No intervention | Government Healthcare Payers | Dialysis Facilities |
| Chang (2023) | Retrospective Cohort | Taiwan | National Level | PD-encouraging reimbursement policy | Historical control | Government Healthcare Payers | Dialysis Facilities |
| Johansen (2023) | Retrospective Cohort | USA | National Level | End-Stage Renal Disease Treatment Choices (ETC) Payment Model | Historical control | Government Healthcare Payers | Dialysis Facilities |

The interventions are classified into three main groups: education, service provision, and payment. The most common intervention types among the included studies were education (17 of 43; 40%)^24–40^ followed by service provision (12 of 43; 30%)^24,41,43–52^, and payment (11 of 43; 26%)^56–66^. Additionally, three studies assessed the effect of combined education with service-provision interventions (3 of 43; 7%)^53–55^.

All educational and service-related interventions were provided by nephrologists and nurses, or a multidisciplinary care team consisting of a combination of nephrologists and nurses, together with other relevant professionals, such as renal dieticians, trained kidney educators, social workers, pharmacists, and psychologists. In one study, the educational interventions were also led by existing patients who had the experience of undertaking HoD^24^, and in another study, the educational intervention was led by a government healthcare payer (i.e., Medicare)^36^. For payment, the majority of these interventions were led by the government, except for one study^62^, which examined the impact of private insurance, where patients themselves paid for the insurance, on HoD utilization.

The reported outcomes focused on the initiation and utilization of PD/HoD, and HHD. PD/HoD initiation refers to the number of CKD patients who started PD/HoD as their first dialysis option, while PD/HoD utilization refers to the number of CKD patients currently using PD/HoD at the time of outcome measurement. HHD initiation and HHD utilization were reported in the same manner. Outcomes related to HoD dialysis retention were only reported for PD but not for HHD. PD retention is defined as the number of PD patients who did not switch to ICHD or KT. In studies where the PD drop-off or technique failure rates were reported, the inverse was calculated to express the outcomes homogenously as PD retention to facilitate comparison between studies.

### Education

Out of 17 studies evaluating the effectiveness of education, seven studies reported outcomes related to the initiation of PD/HoD, and three studies focused on PD/HoD utilization outcomes. Three studies measured both the initiation and utilization of PD/HoD. Two studies reported on both PD and HHD utilization. Additionally, one study reported on PD retention, and one study covered both PD initiation and PD retention outcomes.

Educational interventions for CKD patients primarily aim to equip them with the knowledge necessary to navigate KRT options. These programs provided comprehensive information on KRT, covering dialysis techniques and the advantages and disadvantages of each option. Education was delivered by a multidisciplinary care team—including nurses and experienced PD patients—to offer varied perspectives. A range of teaching methods, such as face-to-face sessions, simulation-based teaching, videos, and web-based platforms, were used to improve patient engagement and understanding. Ultimately, these programs supported patients in making informed, collaborative decisions with their dialysis team regarding the best KRT method for their individual needs. The effectiveness of educational interventions from each study is presented in Table 2.

**Table 2.**
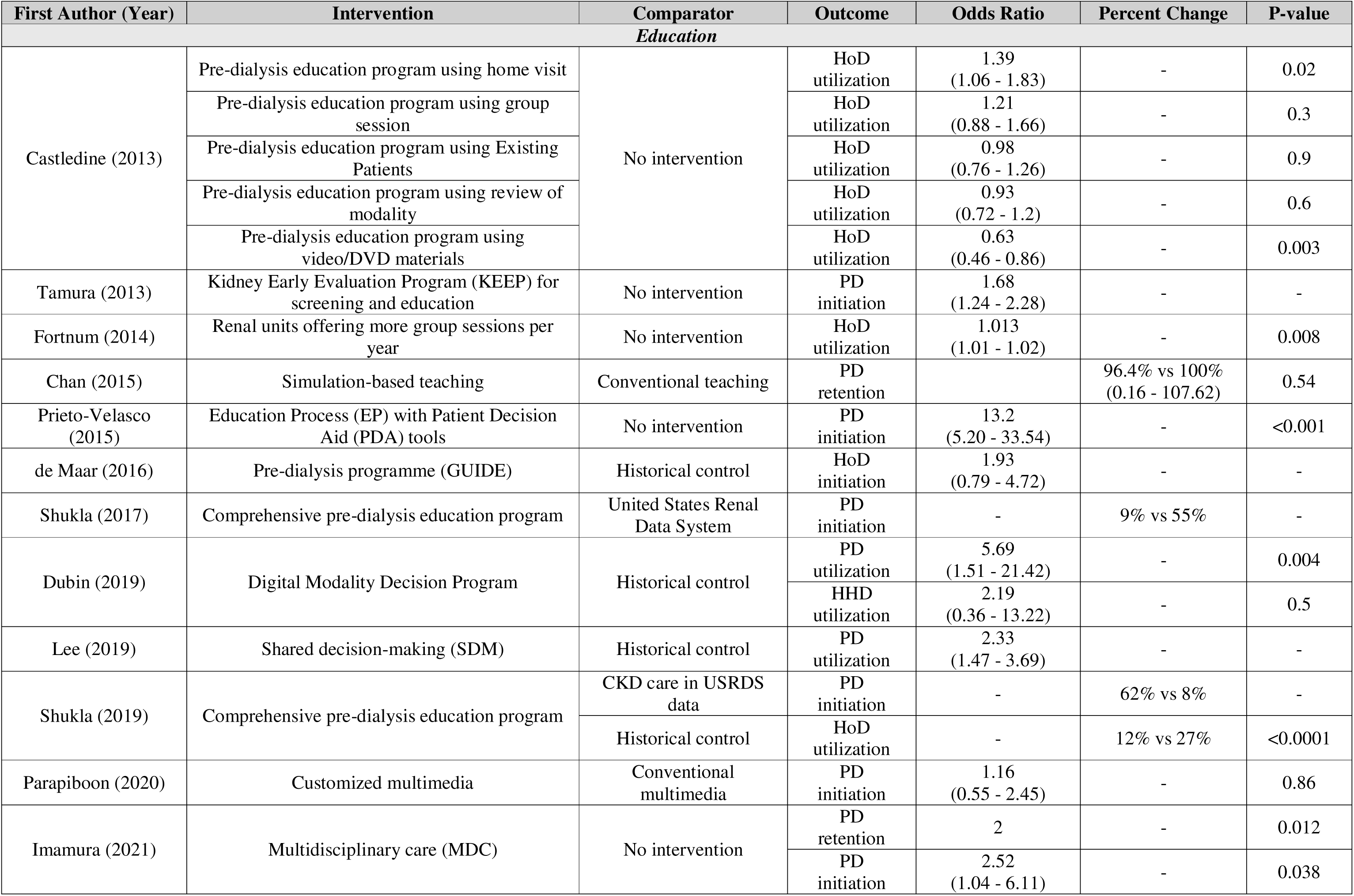
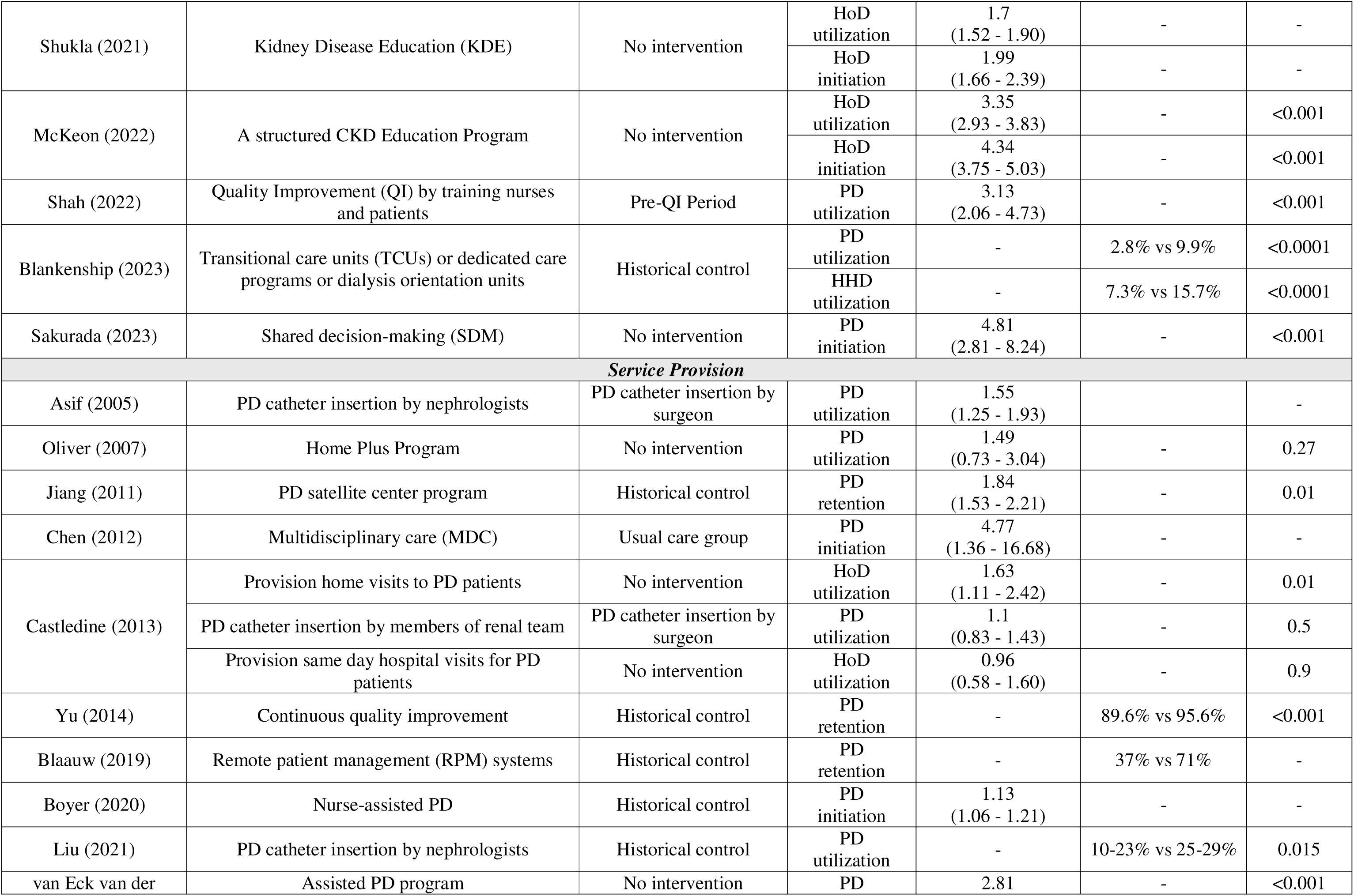
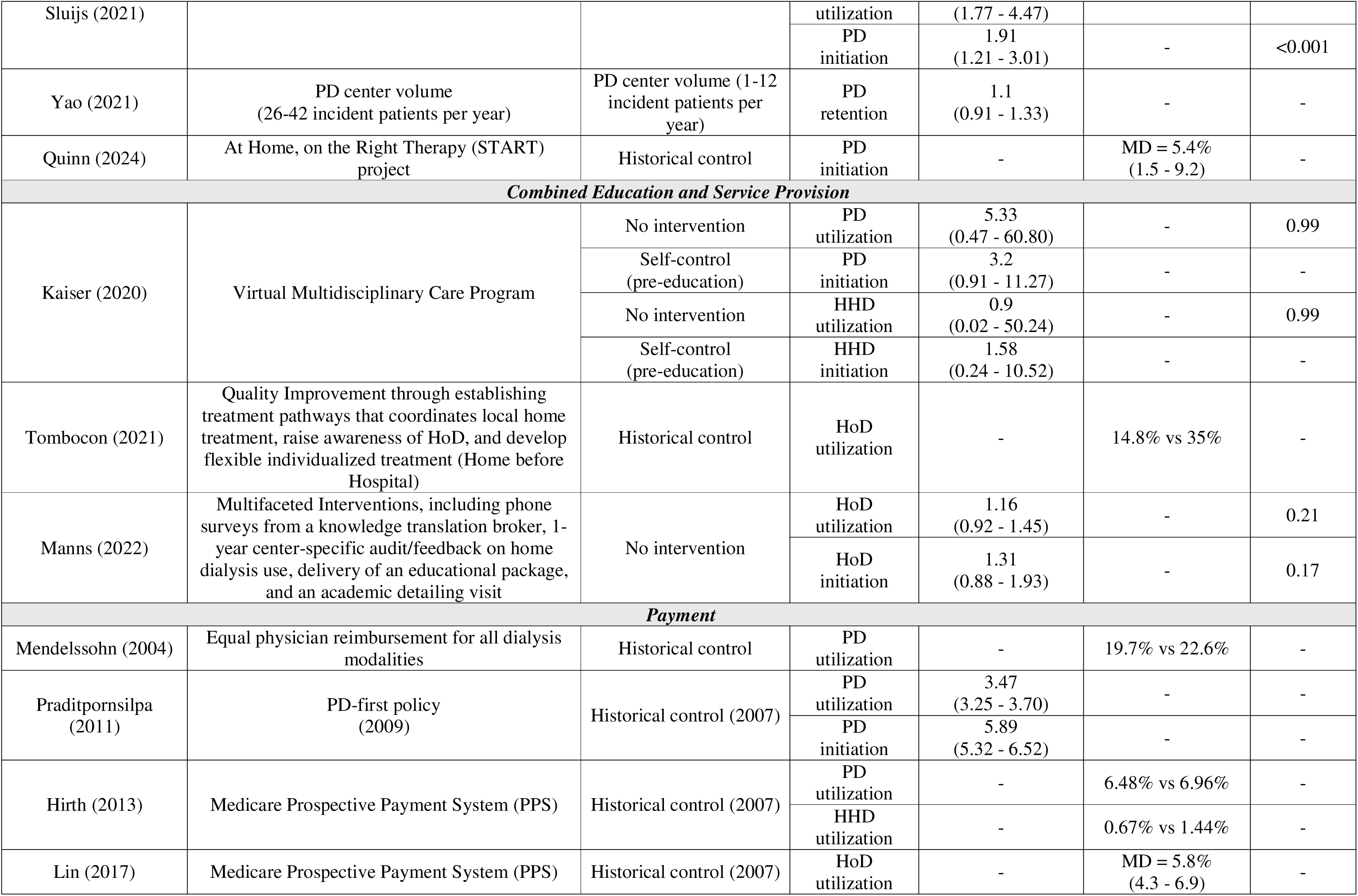
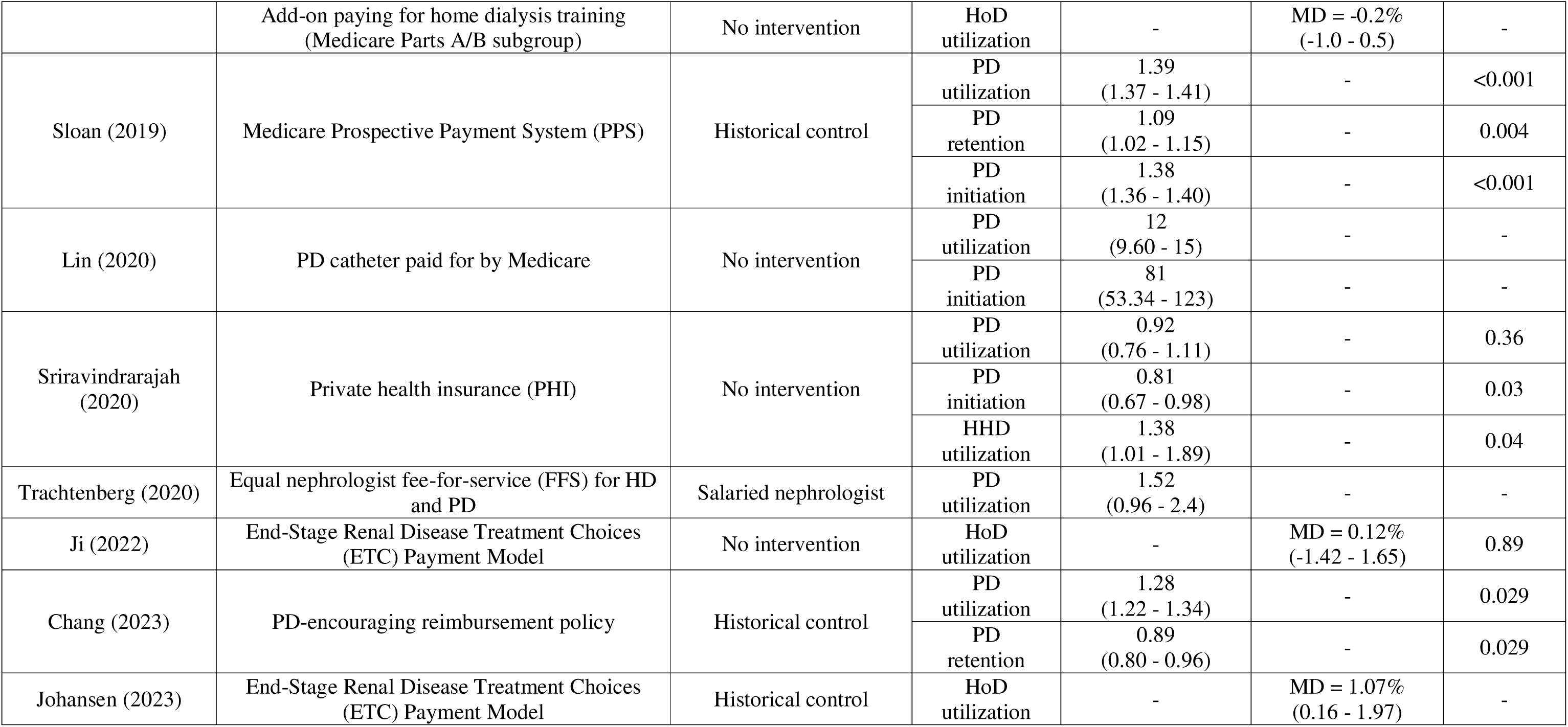
Results of Included Studies.

Regarding the outcome of PD/HoD initiation^25,28–30,33–37,40^, six of the ten studies reporting this outcome found that providing education about PD/HoD significantly increased the initiation of PD/HoD^25,28,35–37,40^. The remaining four studies also observed an increase in PD/HoD initiation in patients receiving this intervention, but these effects were not statistically significant^29,30,33,34^. Additionally, two studies^27,35^ evaluated the outcome of PD retention, with all of them showing that educational interventions increased the retention rate of PD, although only one study reached statistical significance in this regard^35^.

Nine studies reported outcomes on the utilization of PD/HoD^24,26,31–33,36–39^. All of these studies found that educational interventions significantly increased the utilization of PD/HoD compared to no intervention. However, Castledine et al.^24^ found impact varied according to the modality of education delivery (e.g., via home visits, group sessions, video materials, and patients having the experience of performing PD). Specifically, they found that among the education delivery methods investigated, only providing education intervention using home visits significantly increased the rate of PD/HoD utilization in dialysis-requiring patients.

For HHD utilization, results were conflicting between the two studies reporting this outcome. Findings from Blankenship et al. demonstrated a significant benefit of educational interventions in increasing HHD utilization, while results from Dubin et al. found a non-significant benefit of educational intervention in increasing HHD utilization ^31,39^.

### Service Provision

Among the 12 studies evaluating the effectiveness of service provision interventions, three studies reported on the initiation of PD/HoD, while four studies focused on the utilization of PD/HoD outcomes. One study measured both PD/HoD initiation and utilization, and four studies assessed the PD retention rate.

In contrast to educational interventions which focus on pre-dialysis and support the decision-making process, service provision interventions are aimed at enhancing the delivery of dialysis care. These interventions included assisted PD where nurse or other health care providers help patients perform PD at home, catheter insertion performed by nephrologists rather than surgeons, and quality improvement programs, which often involved a multidisciplinary care team.

Regarding four studies reporting the outcome of PD/HoD initiation, all of which involved interventions such as assisted PD and improving PD care quality through a multidisciplinary care team^45,48,50,52^. These studies found that service provision interventions significantly increased the rate of PD initiation compared to no intervention.

For four studies reporting on PD retention outcomes, each assessing the impact of improving the quality of care using different techniques^44,46,47,51^. Two studies provided closely integrated services between hospital and home, and both found that this approach significantly helped patients continue using PD^44,46^. Another study employed telehealth to support patients in performing dialysis at home, which resulted in an increased rate of PD retention^47^. The fourth study evaluated the impact of increasing centre volume on PD retention; however, the study found no significant difference in retention rates between large and small centre volumes^51^.

Among the five studies focused on the outcome of PD/HoD utilization, two studies^43,50^ investigated the effect of home care-assisted PD, and three studies^24,41,49^ assessed the impact of catheter insertion by nephrologists. Of the two studies assessing home care-assisted PD, one found a significant benefit in increasing PD utilization^50^, while one found no significant effect^43^. The results concerning catheter insertion by nephrologists were also inconsistent: two studies reported a significant increase in PD utilization^41,49^, while another found no significant benefit from this intervention^24^.

### Combined Education and Service Provision

Among the three studies evaluating the impact of combined education and service provision interventions^53–55^, one study assessed PD/HoD utilization outcomes^54^, while another examined both PD/HoD initiation and utilization^55^. The third study reported on PD/HoD initiation and utilization as well as HHD initiation and utilization^53^.

Two studies^53,55^ that reported on PD/HoD initiation outcomes observed an increase in initiation rates among patients receiving the combined interventions, though this benefit was not statistically significant^55^. All three studies that evaluated PD/HoD utilization outcomes^53–55^ consistently showed an increase in PD/HoD utilization rates with combined interventions; however, this effect did not reach statistical significance in any of the studies. For the study reporting HHD initiation and utilization outcomes, this study found no significant benefit from the combined interventions in increasing HHD initiation or utilization rates^53^.

### Payment

Of the eleven studies assessing the effectiveness of payment interventions^56–66^, five studies reported outcomes related to PD/HoD utilization^56,59,63,64,66^, while two studies examined both PD/HoD initiation and utilization^57,61^. One study^60^ assessed outcomes for PD/HoD initiation, utilization, and PD retention, and another study^65^ focused on PD/HoD utilization and PD retention. Additionally, one study measured outcomes for PD/HoD initiation, utilization, and HHD utilization^62^.

Payment-related interventions include bundled payments (e.g., Medicare Prospective Payment System, henceforth Medicare PPS^58–60^), capitation (e.g., Thailand’s PD-First policy^57^), fee-for-service (e.g., physician fee in Canada^56,63^), pay-for-performance (e.g., End-Stage Renal Disease Treatment Choices Model, henceforth ETC model^64,66^), and private payments (e.g., private health insurance^62^).

Among the four studies reporting outcomes in terms of PD initiation, two studies investigating the impact of the Medicare scheme in the US–specifically, the Medicare PPS and coverage for PD catheters, found a significant increase in PD initiation following the implementation of these payment interventions^60,61^. Thailand’s PD-First policy also led to a statistically significant rise in PD initiation^57^. In Australia, however, access to private health insurance was associated with a lower likelihood of PD initiation, and this effect was statistically significant^62^. Two studies reported on PD retention with inconsistent results^60,65^. Sloan et al., investigating, found that the payment system with the US Medicare PPS was associated with higher rates of PD retention^60^. On the other hand, Chang found that Taiwan’s PD-encouraging reimbursement policy was associated with lower PD retention rates^65^.

PD/HoD utilization was reported in eleven studies^56–66^. Interventions that were associated with a significant increase in PD/HoD utilization were the US Medicare PPS, Taiwan’s PD-encouraging reimbursement policy, and Thailand’s PD-First policy^57,59–61,65^. However, Medicare’s home dialysis training add-on was not associated with a significant increase in PD/HoD utilization^59^. Mixed results were found for the ETC model: two studies^64,66^ found that the model was associated with an increase in HoD utilization, but the impact was only statistically significant in one study^66^. The patient having supplementary private health insurance in Australia^62^ and the increase of the PD fee-for-service for nephrologists to be equivalent to HD^63^ in Canada were not associated with significant increases in PD utilization.

Regarding the impact of payment interventions on HHD utilization^58,62^, one study^62^ found that providing supplementary private health insurance significantly increased HHD utilization. However, the implementation of the Medicare Prospective Payment System (PPS) did not result in an increased HHD utilization rate.

## DISCUSSION

This scoping review provides a comprehensive analysis of public health interventions aimed at enhancing the initiation, utilization, and retention of HoD, including both PD and HHD. Our findings indicate that education and service provision interventions can effectively increase initiation, utilization, and retention rates of HoD in patients requiring dialysis, with benefits observed across various types of these interventions. However, the impact of payment interventions on HoD initiation, utilization, and retention varied, showing inconsistent effects depending on the specific type of payment intervention^60,63^.

The decision-making process for selecting a dialysis modality is complex and involves balancing multiple factors, including physician expertise and practices, patient and family values, and the patient’s autonomy and self-management capability^67^. This complexity contributes to the low utilization of PD, despite previous evidence showing that patients on PD and ICHD experience similar mortality outcomes^68,69^. Barriers to HoD utilization can be categorized as those impacting patients—such as limited knowledge, lack of social support, and living in remote areas—as well as barriers within healthcare providers (e.g., reimbursement issues) and the healthcare system (e.g., limited PD catheter access and late referrals to nephrologists^70^). Addressing these barriers through pre-dialysis education, adjustments in service provision, and modifications to payment structures may increase HoD utilization among dialysis-requiring patients.

Our review found that most of the studies assessing the effectiveness of educational interventions show a significant benefit in increasing the utilization and retention of HoD in dialysis-requiring patients. Successful educational programs often stemmed from the pre-dialysis education initiatives that provided comprehensive information on KRT options. To illustrate, healthcare providers may help patients through an exercise where they draw out how different dialysis modalities may be incorporated into their weekly timetable^28^. Additionally, patients may be asked to state the pros and cons of each dialysis modality and assign weights to each factor based on their personal preference^28^. Beginning this process well in advance of when patients require dialysis ensures ample time for shared decision-making among patients, families, and healthcare providers^24–26,28,31–33,36,37,40^. In addition, nearly half of the educational interventions that achieved statistically significant increases in the utilization and retention of HoD were led by multidisciplinary care teams^28,31,32,35,37^. These findings emphasize the importance of incorporating multidisciplinary personnel in improving the effectiveness of the interventions.

The mode of education delivery also plays a critical role; for example, the results from Castledine et al. suggest that providing education via home visits has proven more effective than providing video-based education^24^. Therefore, further investigation into the specific benefits of different educational delivery methods is necessary to draw more meaningful conclusions.

Service provision interventions included assisted PD, which enables patients to perform PD at home with support from nurses or a multidisciplinary care team. Other service provision interventions involved having nephrologists, rather than surgeons, handle PD catheter insertions and implementing mobile or telehealth systems to monitor and assist patients in managing HoD. Our study found that most studies evaluating these approaches reported significant benefits in increasing HoD initiation and utilization rates, especially through assisted PD and catheter insertions performed by nephrologists. A possible explanation for the increased PD uptake rates when nephrologists handle catheter insertions is the reduced delay in starting PD. When surgeons manage catheter insertions, scheduling challenges, and the prioritization of emergency cases often result in delayed PD initiation^71–73^, especially when patients need to be referred to a different healthcare facility to undergo this procedure.

Our study indicates that assisted PD can enhance the utilization and retention of HoD, especially among elderly and physically dependent patients requiring dialysis^74^. These patients often face distinctive obstacles to self-managed dialysis, including a higher prevalence of comorbidities compared to younger patients and a loss of independence due to increasing frailty, which leads to a greater need for caregiver assistance. Providing an assisted PD program for these individuals could be an effective approach to increasing PD use within this group.

Unlike educational and service provision interventions, which show consistent results across various interventions in the same group, the effectiveness of payment interventions found in our review varied depending on the specific type of payment intervention used as well as the context of the health system in which the policy was applied. Illustratively, the 2008 PD-First policy in Thailand was the payment intervention demonstrating the highest impact, with an OR of 5.89 for PD initiation and 3.47 for PD utilization^57^. This significant impact arose from making dialysis services accessible to previously underserved populations and designating PD as the first line of treatment. Conversely, an initiative to promote home dialysis among patients already accessing other forms of dialysis did not achieve similar success: raising nephrologist fee-for-service to match HD fees in Canada, where national health insurance covers both PD and HD services, did not lead to significant change in PD usage^63^.

In contexts where the cost of PD provision is lower than that of ICHD, such as the US and Taiwan, bundled payments were successful at increasing HoD usage^15,75,76^. Studies showed that the Medicare PPS correlated with a significant increase in HoD use, although this effect was not statistically significant for the training add-on^58–60^. Taiwan’s bundled payment, subject to a global budget, has been effective in increasing PD utilization, yet it has also led to a lower PD retention rate^65^. The odds of PD drop-off were 1.33 times higher in clinics compared to medical-centre hospitals, suggesting that inadequate medical knowledge may contribute to reduced retention^65^.

Interestingly, the relationship between private health insurance and home dialysis modality utilization revealed that supplementary private health insurance we associated with higher odds of HHD utilisation but lower odds of PD initiation^62^. However, this study did not control for income as a confounder; those who can afford private health insurance are often better off financially and may be more likely to utilize HHD due to better living conditions^62^.

Overall, education, service and payment-related interventions can contribute to higher home dialysis initiation, retention, and utilization. However, only three studies^53–55^ investigated interventions in more than one of these three groupings. Therefore, the synergetic effects of these interventions could not be clearly understood. Additionally, public health interventions to increase home dialysis usage may be achieved more than via education, service provision, or payment, for example through amending regulations or legislation^77,78^, but their effectiveness are not assessed in the literature. For example, while the Advancing American Kidney Health Executive Order^79^ explicitly supports the use of HoD, we did not find any studies which examines its impact–likely due to the technical difficulties associated with quantitatively assessing high-level interventions such as an executive order. Nevertheless, studies assessing the impact of the End-Stage Renal Disease Treatment Choices (ETC) payment model, which arose as a result of the executive order, were included in our review^64,66^.

Our scoping review has several strengths. Firstly, we provide a comprehensive review of the effectiveness of various public health interventions on the initiation, utilization, and retention of HoD. Additionally, we considered both PD and HHD as outcomes of interest. The evidence on HHD utilization offers valuable insights, as lessons learned from HHD provision in HICs may also apply to PD provision in LMICs.

However, our study has some limitations. A key limitation is the inconsistency in measures of intervention effects, which complicates comparisons of intervention effectiveness across studies. Additionally, most included studies were observational studies and used pre-intervention data as historical controls, which may introduce confounding bias; therefore, further studies with rigorous methodologies are needed to confirm our findings. In addition, our review did not include studies from grey literature, which may lead to publication bias in our findings. Lastly, the studies included in this review were primarily conducted in HICs. This focus underscores a significant gap in evidence from resource-limited settings.

In conclusion, this scoping review suggests that enhancing education and service provision may be the most effective public health strategies for improving initiation, utilization, and retention rates of HoD among dialysis-requiring patients. These findings provide valuable insights for prioritizing policy interventions to support the initiation, uptake, and sustained use of home dialysis both in Thailand and globally.

The findings from this scoping review were presented to a dialysis policy working group and the results were submitted as policy recommendations to the National Health Security Office (NHSO)–the government body managing Thailand’s Universal Health Coverage program. Looking ahead, future research should focus on evaluating these recommended interventions to systematically assess their impacts on dialysis policy.

## METHODS

This scoping was conducted and reported according to the PRISMA-ScR (PRISMA extension for Scoping Reviews)^80^.

### Study identification

Relevant studies were identified through a comprehensive search of six databases including Web of Science, PubMed, Embase, Scopus, EBSCOhost, and EconLit since their inception through May 2024. The search terms used consisted of three domains: *Increase* AND *Utilization* AND *Home Dialysis*. The search terms and search strategies used for each database are shown in Supplementary Table 1. Additionally, the reference lists of the included studies were examined to further identify relevant studies for the review.

### Study selection

The study selection process was facilitated by the Covidence systematic review software (version 2, Veritas Health Innovation, Melbourne, VIC, Australia). Titles and abstracts of the identified studies were screened by one reviewer (all authors). Full texts of the studies were reviewed independently by two reviewers if the decision could not be made based on titles and abstracts (all authors).

Observational studies (i.e., case-control, cross-sectional, and cohort studies), quasi-experimental studies, and randomized controlled trials (RCT) were eligible for this review if they met all of the following criteria: 1) studies that included participants as non-dialysis dependent CKD or dialysis-requiring CKD, and 2) studies that assessed and reported the effect of public health intervention on increasing utilization or retention of HoD. Therapeutic interventions, such as the use of innovative dialysate, were deemed beyond the scope of the review and were excluded.

In this review, “home dialysis” is defined as any dialysis modality conducted at the patient’s house, including PD and HHD. Public health interventions in this review are defined as the interventions that are focused on individual, or system levels^78^. The interventions focused on individual levels aim to change beliefs, attitudes, and knowledge about home dialysis in CKD patients such as education and shared decision-making. The interventions focused on system levels and aimed to change the organization, laws, and policy of home dialysis such as change in service provision (e.g., home visit by nurse, insert catheter by nephrologist), or change in payment system or policy.

### Data extraction

After the study selection process was completed, the included studies then went through a data extraction process by a single reviewer using Microsoft Excel. During this process, data regarding the study characteristics, details of the intervention, study context, impact, costs of implementing the intervention, as well as the supporting and limiting factors to the success of the intervention were extracted. Later, the impact data extracted was then cross-checked by another reviewer (TA and PS).

### Data analysis

The effects of interventions on the utilization and retention of home dialysis were summarized qualitatively by intervention types and outcomes. However, as PD is the predominant home dialysis modality, the term HoD in the studies that did not specify PD or HHD was assumed to refer to PD in our analysis.

## ACKNOWLEDGEMENTS

We would like to extend our gratitude to all parties involved in the Commission: The Learning Committee, chaired by Prof. Emeritus Kriang Tungsanga. Prof. Vivekanand Jha; The National Health Security Office’s Working Group, chaired by Prof. Kearkiat Praditpornsilpa; and the Secretariat team of both the Committee and the Working Group. Their invaluable guidance and feedback have been instrumental throughout the research.

In addition, we would like to thank Kinanti Khansa Chavarina of the Health Intervention and Technology Assessment Program (HITAP) for helping design the first stages of the study and for giving a detailed review of the manuscript. We would also like to thank Siriyakorn Khamkom of the Faculty of Pharmacy, Silpakorn University for her contribution to the title and abstract screening process.

## AUTHOR CONTRIBUTIONS

N.Y., Y.T., S.D., and T.A. contributed to study’s concept and design. N.G. and T.A. searched for the relevant studies. N.Y., Y.T., P.S., D.P., T.C., J.S., N.G., N.K., S.D. and T.A. screened studies for the titles, abstracts, and full texts and performed the data extraction. N.Y., Y.T., and T.A. analysed the data and interpreted the results. N.Y. and T.A. drafted the manuscript. Y.T., P.S., D.P., T.C., J.S., N.G., N.K., and S.D. critically revised the manuscript. All authors approved the final version of the manuscript.

## DATA AVAILABILITY

Not applicable. This scoping review did not apply meta-analysis and only qualitatively summarized the data presented in the primary studies.

