## Supplementary Table for "The Way Home: A Scoping Review of Public Health Strategies to Increase the Utilization of Home Dialysis in Chronic Kidney Disease Patients"

**SUPPLEMENTARY INFORMATION**

**Supplementary Table 1 Search Terms by Database**

| **Database** | **Search Strategy** | **No. of Entries** |
| --- | --- | --- |
| Web of Science | (((TS=(Peritoneal dialysis OR home AND Dialysis )) AND TS=((increase) OR (increasing) OR (enhance) OR (enhancing) OR (higher) OR (assisting) OR (assistance) OR (success) OR (successful) OR (initiate) OR (initiating) OR (initiation) )) AND TS=((use) OR (utilizing) OR (utilization) OR (uptake) OR (accept))) | 9,829 |
| PubMed | (((increase OR increasing OR enhance OR enhancing OR higher OR assisting OR assistance OR success OR successful OR initiate OR initiating OR initiation) AND (use OR utilizing OR utilization OR uptake OR accept)) AND (peritoneal OR home)) AND (dialysis OR peritoneal dialysis [MeSH]) Sort by: First Author | 7,752 |
| EMBASE (Ovid) | ('peritoneal dialysis'/exp OR 'dialysis, peritoneal' OR 'intermittent peritoneal dialysis' OR 'peritoneal dialysis' OR 'peritoneal dialysis, intermittent' OR 'peritoneal irrigation' OR 'home dialysis') AND (increasing OR increase OR enhance OR enhancing OR higher OR assisting OR assistance OR success OR successful OR initiate OR initiating OR 'initiation'/exp) AND (use OR utilizing OR utilising OR utilization OR utilisation OR uptake OR accept) | 4,628 |
| Scopus | ((TITLE-ABS-KEY("utilizing")) OR (TITLE-ABS-KEY("utilization")) OR (TITLE-ABS-KEY("uptake")) OR (TITLE-ABS-KEY("acceptance")) OR (TITLE-ABS-KEY("accepting")) OR (TITLE-ABS-KEY("accept"))) AND ((TITLE-ABS-KEY("increase")) OR (TITLE-ABS-KEY("increasing")) OR (TITLE-ABS-KEY("enhance")) OR (TITLE-ABS-KEY("enhancing")) OR (TITLE-ABS-KEY("higher")) OR (TITLE-ABS-KEY("assisting")) OR (TITLE-ABS-KEY("assistance")) OR (TITLE-ABS-KEY("success")) OR (TITLE-ABS-KEY("successful")) OR (TITLE-ABS-KEY("initiate")) OR (TITLE-ABS-KEY("initiating")) OR (TITLE-ABS-KEY("initiation"))) AND ((ALL("home dialysis")) OR (ALL("peritoneal dialysis"))) | 1,932 |
| EBSCOhost | (+peritoneal+dialysis+OR+home+dialysis+)+AND+(+increase+OR+increasing+OR+enhance+OR+enhancing+OR+higher+OR+assisting+OR+assistance+OR+success+OR+successful+OR+initiate+OR+initiating+OR+initiation+)+AND+(+use+OR+utilising+OR+utilisation+OR+uptake+OR+accept+) | 907 |
| EconLit | ((Peritoneal dialysis or Home dialysis) and (increase or increasing or enhance or enhancing or higher or assisting or assistance or success or successful or initiate or initiating or initiation or use or utilise or utilising or accept)).af. | 9 |
